# Cohort profile: the Viral load Cohort North-East Lesotho (VICONEL) from 2016 to 2022 – cohort description, test volumes, predictors of viraemia, and the road ahead

**DOI:** 10.1101/2024.03.12.24304025

**Authors:** Jennifer A. Brown, Lipontso Motaboli, Malebanye Lerotholi, Maurus Kohler, Kathrin Hänggi, Moliehi Mokete, Makobefo G. Chakela, Mpho Kao, Mathebe Kopo, Moleboheng Mokebe, Bienvenu L. Nsakala, Blaise Lukau, Irene Ayakaka, Alain Amstutz, Jochen Ehmer, Thomas Klimkait, Tracy R. Glass, Josephine Muhairwe, Frédérique Chammartin, Nadine Tschumi, Niklaus D. Labhardt

## Abstract

**Purpose:** The prospective Viral load Cohort North-East Lesotho (VICONEL) aims to support clinical management and generate scientific evidence to inform HIV care. Specifically, VICONEL allows for monitoring of HIV treatment outcomes and health system performance, encompasses a biobank for further research with routinely collected blood plasma samples of consenting participants, and provides a valuable framework for nested observational and interventional studies.

**Participants:** VICONEL captures routine viral load test results alongside associated demographic and treatment information among people in care for HIV in Lesotho, southern Africa. As of December 2022, it encompasses all viral load testing from 23 healthcare facilities in two districts of Lesotho.

**Findings to date:** From January 2016 to December 2022, 114’838 viral load test results were available for 27,472 participants. At the time of the last viral load test, median age was 42 years (interquartile range [IQR]: 33-53); 17,324 (63%) were adult women, 9,273 (34%) adult men, and 870 (3%) children <15 years (age/sex missing for 5); and median time taking antiretroviral therapy (ART) was 6.0 years (IQR 3.0-9.2). Overall, the proportion of cohort participants with viral suppression to <1,000 copies/mL has continually exceeded 90% and has been above 95% since 2020; however, this proportion has consistently been lower among children. Sex, age category / ART regimen core agent (combined variable), time since ART initiation, and district were independently associated with viraemia.

**Future plans:** VICONEL offers potential for i) further digitalisation and automation of results sharing at the client, facility, and district/national level, ii) integration of additional clinical and diagnostic data, including HIV comorbidities, and iii) embedding randomised trials.

**Strengths and limitations:**

- VICONEL covers all HIV viral load testing from 23 clinics in two districts in Lesotho and is thus highly representative.
- Data capture occurs at the time point of viral load testing; thus, treatment or clinical data are not updated between viral load tests, and reasons for exiting the cohort are not followed up.
- Participant data beyond viral load results are limited to key demographic, clinical, and treatment information.
- The cohort and associated biobank have proven to be a valuable platform for nested observational and interventional research, including randomised trials.
- Core functions can be maintained at low cost, constituting a model for near-real-time monitoring of treatment outcomes with limited resources.

## Introduction

Prospective cohorts provide longitudinal data to monitor services and health outcomes within HIV programmes. This facilitates the identification of gaps in service delivery and uptake, early detection of emerging public health threats, and assessments of temporal trends or effects of guideline changes, potentially informing future guidelines[1,2]. Furthermore, they can serve as a platform for nested randomised clinical trials using the routinely collected data for recruitment and endpoint collection.

Regular viral load testing is essential to monitor the effectiveness of antiretroviral therapy (ART) for people with HIV and to take timely action if required. Indeed, the World Health Organization has recommended viral load testing as the preferred method of monitoring treatment outcomes since 2013[3]. In Lesotho, southern Africa, viral load monitoring began shortly after these recommendations but was initially centralised in the capital city, Maseru. Starting in 2015, a consortium consisting of the Ministry of Health of Lesotho, the District Health Management Team of Butha-Buthe, the non-profit organization SolidarMed, the Swiss Tropical and Public Health Institute, and the University of Basel collaboratively supported the District Laboratory of Butha-Buthe Government Hospital in northern Lesotho to launch the first decentralised viral load testing platform in the country. At the time, access to viral load monitoring in the periphery was severely restricted due to limited capacity for centralised testing and the need for long-distance transport of samples and paper-based results. The decentralised viral load testing laboratory and an associated viral load database were launched in December 2015 and officially inaugurated by the Minister of Health in June 2016. After six months of piloting service provision for Butha-Buthe Government Hospital, viral load testing through this laboratory was gradually rolled out to all clinics in Butha-Buthe district and, from 2018 onwards, to neighbouring Mokhotlong district. In conjunction with building the laboratory infrastructure and database, the Viral load Cohort North-East Lesotho (VICONEL) was established to monitor viral load outcomes among persons with HIV who are in care in Mokhotlong or Butha-Buthe district.

Here, we report on cohort composition, test numbers, treatment outcomes, and factors associated with viraemia from the first seven years of the cohort (2016 to 2022), review the cohort’s broader scientific findings so far, and outline the road ahead.

## Cohort description

### Cohort design

VICONEL was designed in conjunction with the establishment of decentralised viral load testing in Butha-Buthe, Lesotho. The prospective open cohort includes people with HIV who receive viral load monitoring with samples tested at the laboratory of Butha-Buthe Government Hospital or, since 2020, through near-point-of-care viral load monitoring currently available at three participating clinics (Seboche Mission Hospital, Mokhotlong Government Hospital, Mapholaneng Health Centre). It includes people in care in Butha-Buthe district since the start of the cohort, and people in care in Mokhotlong district since 2018. As of December 2022, three hospitals, 18 peripheral nurse-led health centres, one private clinic, and one clinic for mine workers were included in the cohort. The cohort study includes a biobank storing leftover plasma samples from viral load testing.

### Objectives

VICONEL aims to support local clinical management of HIV and generate scientific evidence to inform local and global HIV care. The former is achieved in part through a dashboard for clinicians and automated reports flagging persons with incident or repeated viraemia to facilitate clinical follow-up. Scientific evidence is generated through analysis of routine data[4–8], as well as nested observational[9–12] and interventional studies[13].

### Setting

At 19%, Lesotho has the second-highest adult HIV prevalence among all countries globally[14]. In 2022, 270’000 people in the country were living with HIV, and 4’800 people newly acquired HIV[14]. The majority of the population lives rurally, impeding access to healthcare. This is further compounded by the scarcity of medical professionals, with just 4.7 medical doctors and 32.6 nursing and midwifery professionals per 10’000 population[15].

VICONEL covers two of the ten districts in the country with a combined population of around 220’000 people[16]. Previous analyses within the cohort have demonstrated serious gaps along the viral load care cascade for children and adults with HIV in this setting[4,5].

Beyond the progressive rollout of decentralised viral load testing (and the VICONEL cohort), the rollout of ART containing the integrase strand transfer inhibitor (INSTI) dolutegravir as a core agent has constituted another key change in care provision during the reporting period. Dolutegravir rollout began in Lesotho in late 2019, with the main programmatic transition from non-nucleoside reverse transcriptase inhibitor- (NNRTI-) to dolutegravir-based ART occurring throughout 2020. For children taking ART containing the protease inhibitor ritonavir-boosted lopinavir as the core agent, the main transition period to dolutegravir-based ART began in and continued beyond 2022.

### Procedures

For the period 2016-2022, according to national guidelines, adults with HIV should receive viral load testing six and 12 months after starting ART or switching to second- or third-line ART, as well as annually thereafter, whereas six-monthly testing is recommended for children and adolescents (0-19 years)[17,18]. More frequent viral load testing is indicated during pregnancy and breastfeeding, as well as upon detection of viraemia[17,18].

Blood samples collected in healthcare facilities in Butha-Buthe or Mokhotlong districts are transported to one of the three hospital laboratories, typically by a motorbike transport service, for plasma separation. A national Viral Load Request Form is filled out by the treating facility and transported with the sample. Upon reception at the hospital laboratory, data is entered into the Laboratory Information System (LIS) managed by the Ministry of Health. If the sample is first received in one of the two other hospital laboratories, the separated plasma is then transported to the Butha-Buthe Government Hospital laboratory. Viral load testing was initially performed using a COBAS Ampliprep/COBAS TaqMan HIV-1 Test, v2.0, and later using the COBAS 4800 system HIV-1 Test (F. Hoffmann-La Roche AG, Basel, Switzerland). Samples are linked to the instrument position via barcode scanning before viral load testing. Once the run is complete and the testing laboratory has authorised results, viral load results feed into the LIS, upon which the referring laboratories can review data and paper-based results forms can be printed and distributed to the respective facilities. Data is extracted from the local LIS approximately weekly and uploaded to the VICONEL database maintained by SolidarMed and the Division of Clinical Epidemiology, University Hospital Basel. The data team of SolidarMed Lesotho then confirms each viral load result and associated information, cross-checking the Viral Load Request Form against existing data in the database and updating the database information (e.g., ART regimen, regimen start and stop dates) if applicable. If necessary, healthcare facilities are contacted to complete missing or unclear information. In addition, the VICONEL data manager runs data quality checks on a regular basis and investigates unusual entries. Data routinely collected in the cohort database include demographic, clinical, laboratory, treatment, consenting, and biobank data.

In addition to laboratory-based testing, since 2020, a small proportion of people with HIV have received near-point-of-care viral load testing using Xpert HIV-1 Viral Load on GeneXpert Systems (Cepheid, Sunnyvale, CA, United States), available at two hospitals and one health centre. According to current guidelines, point-of-care viral load testing is prioritised for infants, children, adolescents, for pregnant and breastfeeding people, and people with HIV viraemia[18]. Currently, viral load results from GeneXpert Systems are not integrated into the LIS, necessitating manual collection of associated participant data from Viral Load Request Forms and local registries.

Plasma samples left over after viral load testing are stored in a biobank for clinical and, if informed consent for further use is provided, research purposes. At the time of writing, this is operationally limited to viraemic samples.

The cohort does not interfere with the scheduling of viral load testing. However, its database is used, notably by local clinicians, to support clinical management.

### Analyses and statistical methods

The data presented here includes viral load results from 1 January 2016 to 31 December 2022. We exclude entries i) where the viral load result is missing (i.e., failed tests); ii) where viral load testing occurred before the first-ever ART initiation (not part of routine care in Lesotho); iii) where both the date of blood draw and the date of testing are missing, or iv) from unknown facilities, facilities contributing less than 50 viral load results throughout the reporting period, or from referring facilities outside of Butha-Buthe or Mokhotlong district. If multiple viral load tests are conducted for the same individual on the same day (e.g., due to suspected testing errors), the highest viral load is considered. We include individuals with at least one available viral load result after application of these criteria.

We report on cohort composition, viral load testing and turnaround time (defined here as time from blood draw to viral load testing), and the proportion of participants with viraemia by year. Ages <15 years are defined as children and ages ≥15 years as adults, in line with UNAIDS reporting[14]. In addition, we assessed for factors associated with viraemia ≥50 copies/mL or ≥1,000 copies/mL at the level of the individual viral load result using a mixed effect logistic regression model with the participant as the random effect. For this analysis, age category and ART core agent were represented in a single variable due to collinearity. When the core agent was changed immediately after a documented viral load ≥50 copies/mL or ≥1,000 copies/mL, respectively, viral load results following this regimen change were excluded to avoid misleading interpretation, as switching treatment due to viraemia represents a unique opportunity for resuppression within an individual.

All statistical analyses were performed using Stata MP version 16.1.

## Findings to date

### Participant characteristics over time

Overall, 27,472 individuals living with HIV received at least one valid viral load result through this platform from January 2016 through December 2022 and were thus included (**Table 1**). Of those with at least one viral load result in 2022, 10,679/17,944 (60%) were female, median age was 44 years (IQR 35-54), 619/17,944 (3%) were children <15 years of age, median time since ART initiation was 6.6 years (IQR 3.9-10.2), and the most common ART regimen was tenofovir disoproxil fumarate (TDF) / lamivudine (3TC) / dolutegravir (DTG), prescribed to 15,826/17,944 (88%) participants. The number of individuals receiving at least one viral load test in any given year increased in line with initial cohort expansion, with a particular jump in 2018 as Mokhotlong district was included, and has stabilised since 2020 (**Table 1**). Median follow-up time from the first to the last available viral load result was 3.0 years (IQR 1.0-4.6).

**Table 1:**
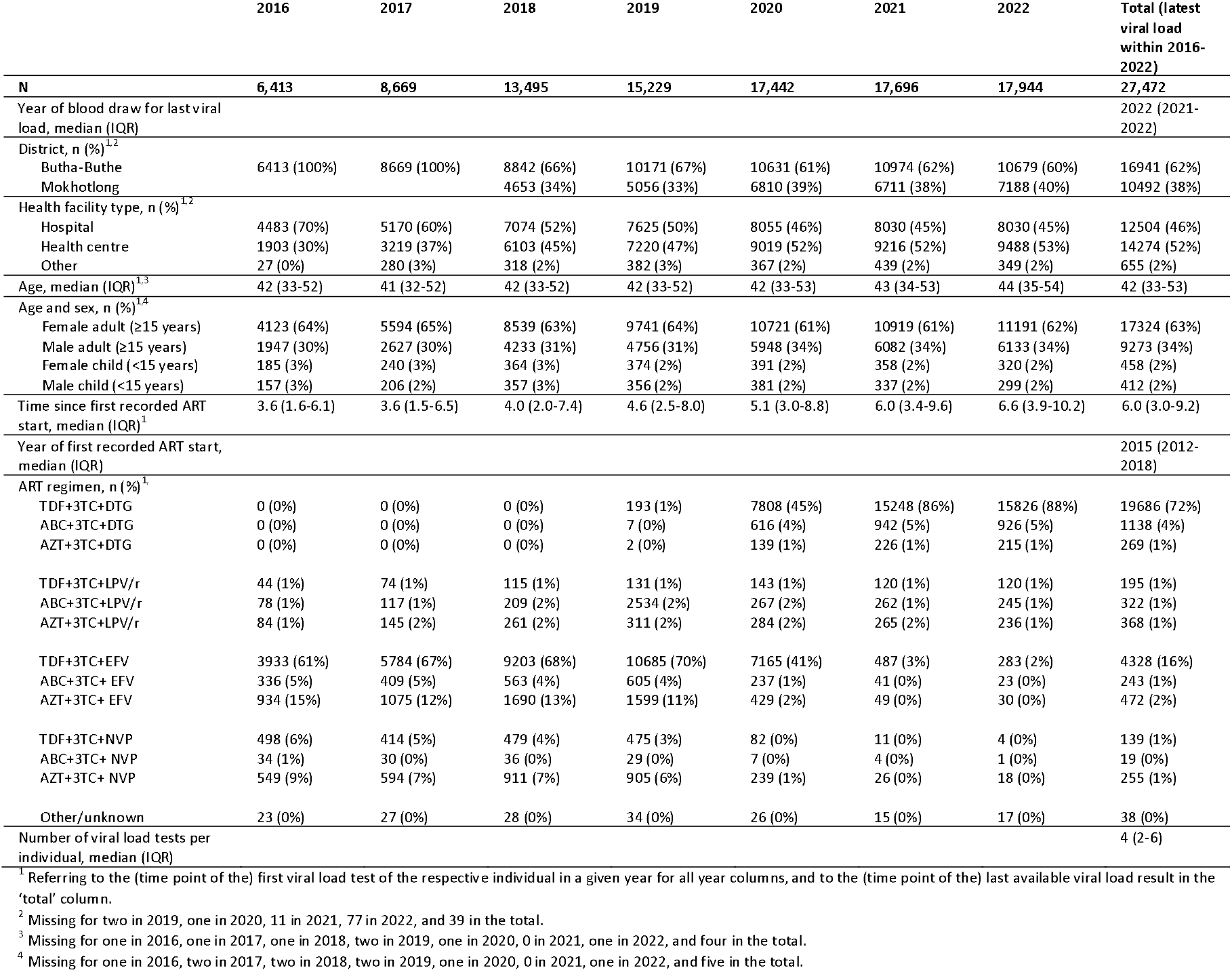
Participant characteristics. For each year, cohort participants with at least one viral load test in the respective year are shown. Data refers to the (time point of the) first viral load test of a given individual in the respective year. In the ‘total’ column, data refers to the (time point of the) last available viral load test for each individual. 3TC: lamivudine; ABC; abacavir; AZT: zidovudine; DTG: dolutegravir; IQR: interquartile range; EFV: efavirenz; LPV/r: ritonavir-boosted lopinavir; NVP: nevirapine; TDF: tenofovir disoproxil fumarate.

### Viral load testing over time

From 1 January 2016 to 31 December 2022, 114,838 viral load tests were conducted. The annual number of samples increased each year until 2020, followed by a slight decrease and stabilisation (**Figure 1A**). The time between blood draw and laboratory-based testing varied greatly by year but overall decreased and stabilised from 2020 to 2022, with a median of 5 (IQR 2-7) days in 2022 (**Figure 1B**). While we did not specifically assess the effect of the COVID-19 pandemic on viral load testing, major disruptions in viral load testing during the pandemic as reported for other settings[19] are not immediately apparent for this cohort.

**Figure 1:**
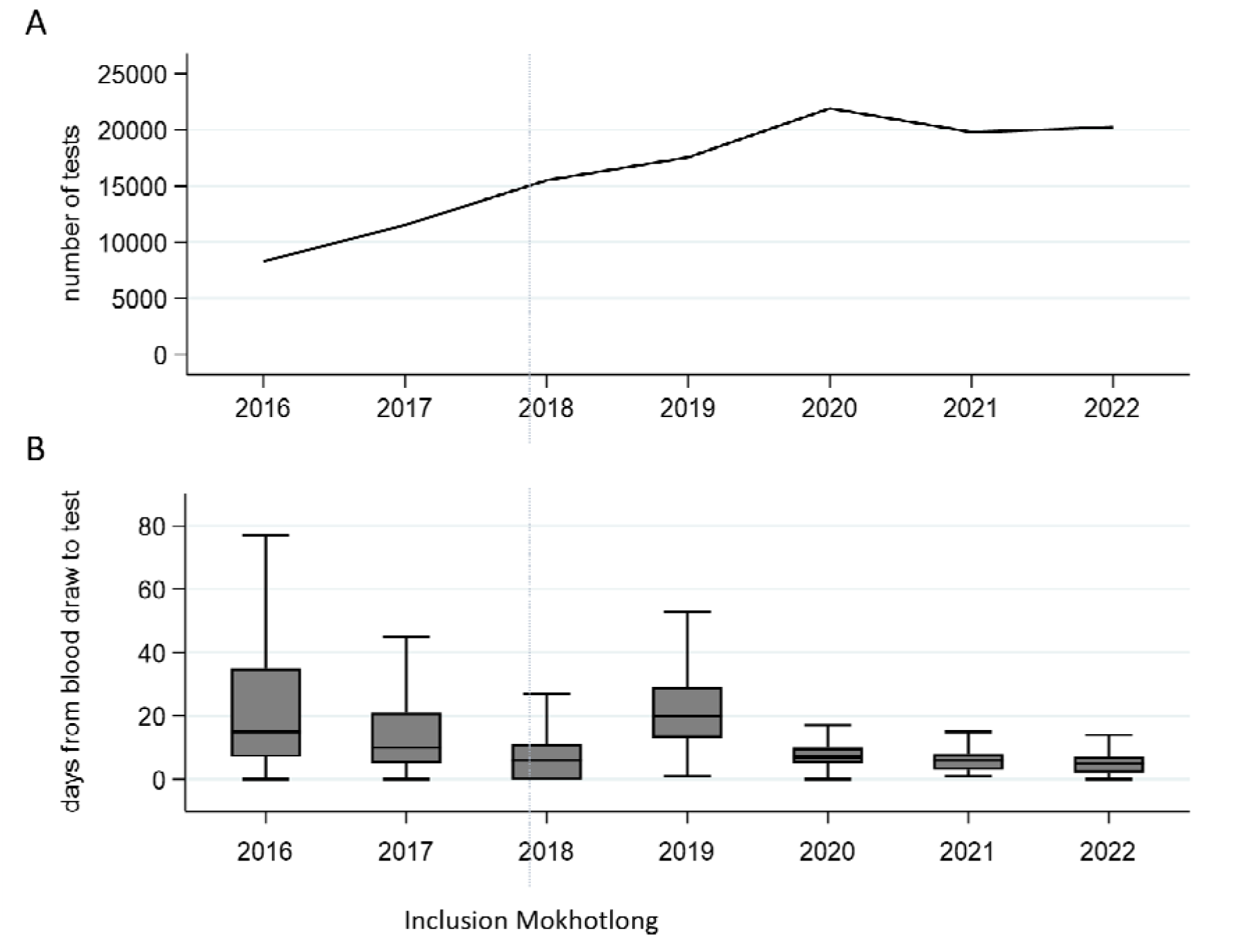
Viral load testing over time. A: Number of viral load tests by year. B: Box plots showing turnaround time, defined as the number of days from blood draw to viral load testing, by year (outside values are excluded).

### Virological outcomes over time

Viral suppression rates to <1,000 copies/mL have consistently exceeded 90% since the launch of the cohort, and have exceeded 95% since 2020 (**Figure 2**). Increases in the proportion of participants with viraemia in 2018/2019 might be partially explained by the inclusion of Mokhotlong district from 2018 onwards, as the rate of viraemia has consistently been higher in Mokhotlong than in Butha-Buthe district (**Figures S1-S2**). However, a slight increase was also observed in Butha-Buthe, pointing towards additional underlying factors. The subsequent decreased prevalence of viraemia from 2020 onwards appears temporally aligned with the rollout of dolutegravir in Lesotho. Throughout the observation period, viral suppression rates among children have been substantially lower than among adults; this is consistent with reported challenges in achieving viral suppression among children and global reports[14,20].

**Figure 2:**
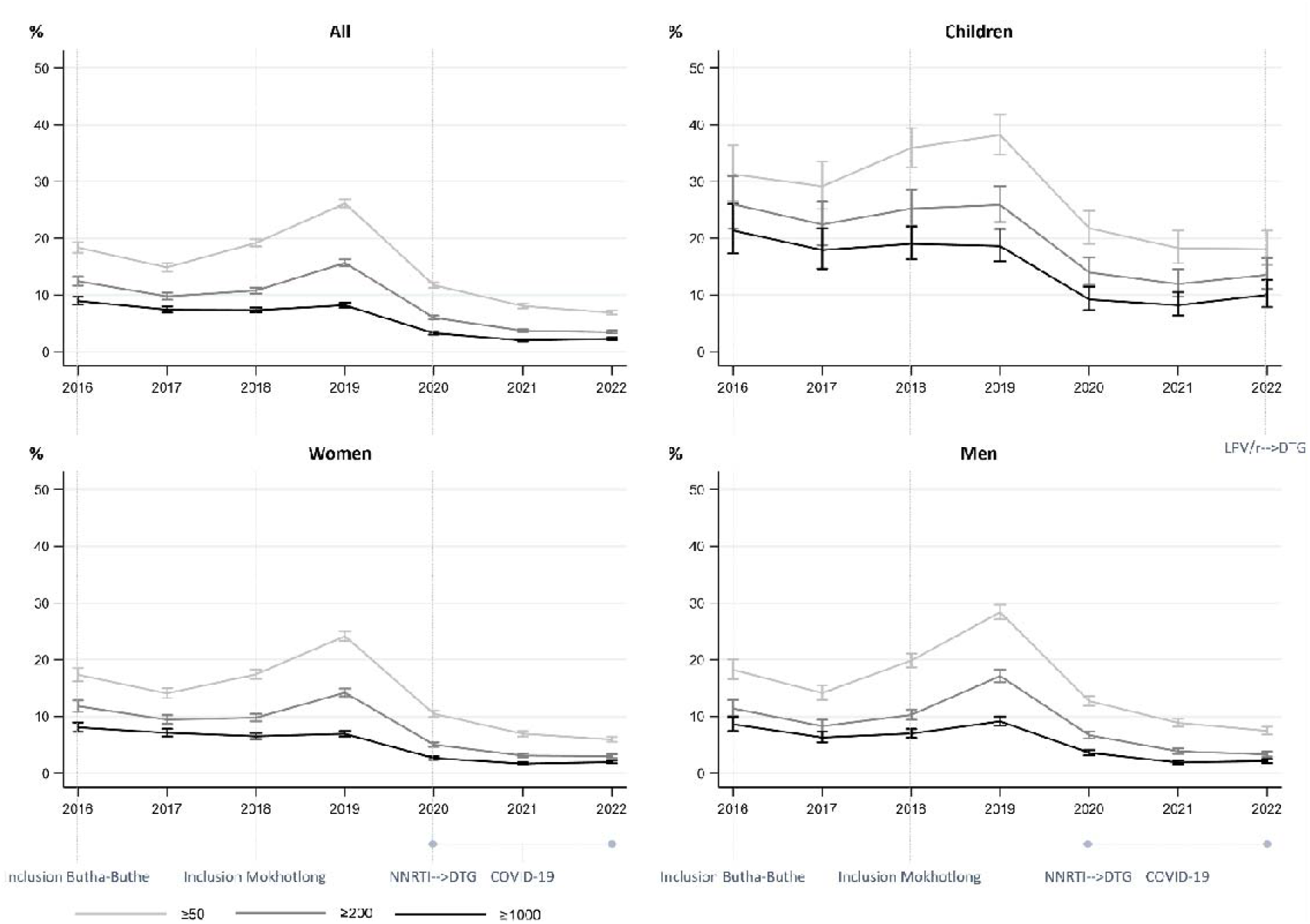
Rates of viraemia above various thresholds (in copies/mL) over time. The first viral load result of any individual in a given year is considered. The denominator corresponds to the number of individuals receiving at least one viral load test in a given year. Bars indicate 95% confidence intervals. DTG: dolutegravir; LPV/r: ritonavir-boosted lopinavir; NNRTI: non-nucleoside reverse transcriptase inhibitor.

Among participants with at least one viral load test in 2022, at their first test in the year, 415/17,944 (2%) had a viral load ≥1,000 copies/mL and 1,237/17,944 (7%) had a viral load ≥50 copies/mL. Among adult women, adult men, and children, 221/11,191 (2%), 132/6,133 (2%), and 62/619 (10%), respectively, had viraemia ≥1,000 copies/mL and 666/11,191 (6%), 459/6,133 (7%), and 112/619 (18%) had viraemia ≥50 copies/mL, respectively.

### Factors associated with viraemia

We assessed factors associated with viraemia using a mixed effects logistical regression model among included viral load results. Being female (vs male; adjusted odds ratio [aOR] 0.80, 95% CI 0.75-0.85), increasing time since ART initiation (per five years; aOR 0.94, 95% CI 0.90-0.98), being in care in Butha-Buthe district (vs Mokhotlong; aOR: 0.50, 95% CI: 0.47-0.53) and, compared with adults taking NNRTI-based ART, being an adult taking INSTI-based ART (aOR 0.32; CI 0.30-0.33) or a child taking INSTI-based ART (aOR 0.78; 95% CI 0.62-0.99) were associated with a lower aOR of viraemia ≥50 copies/mL. Being an adult taking PI-based ART (aOR 1.52, 95% CI 1.22-1.88), a child taking NNRTI-based ART (aOR 2.15; 95% CI 1.85-2.50), or a child taking PI-based ART (aOR 2.06, 95% CI 1.65-2.58) were associated with a higher aOR (**Figure 3**). The higher aORs for viraemia among children compared with adults taking the corresponding ART regimen aligns with higher overall rates of viraemia among children. Higher odds ratios of viraemia in the context of protease inhibitors, especially among adults, may be partly related to their use in second-line ART. Lower odds of viraemia with increasing time since ART initiation was still observed when restricting analysis to either children or adults, though the effect was greater among children; furthermore, restricting analysis to children, no effect of sex was observed (for female [vs male] children: aOR 0.95; 95% CI: 0.71-1.28).

**Figure 3:**
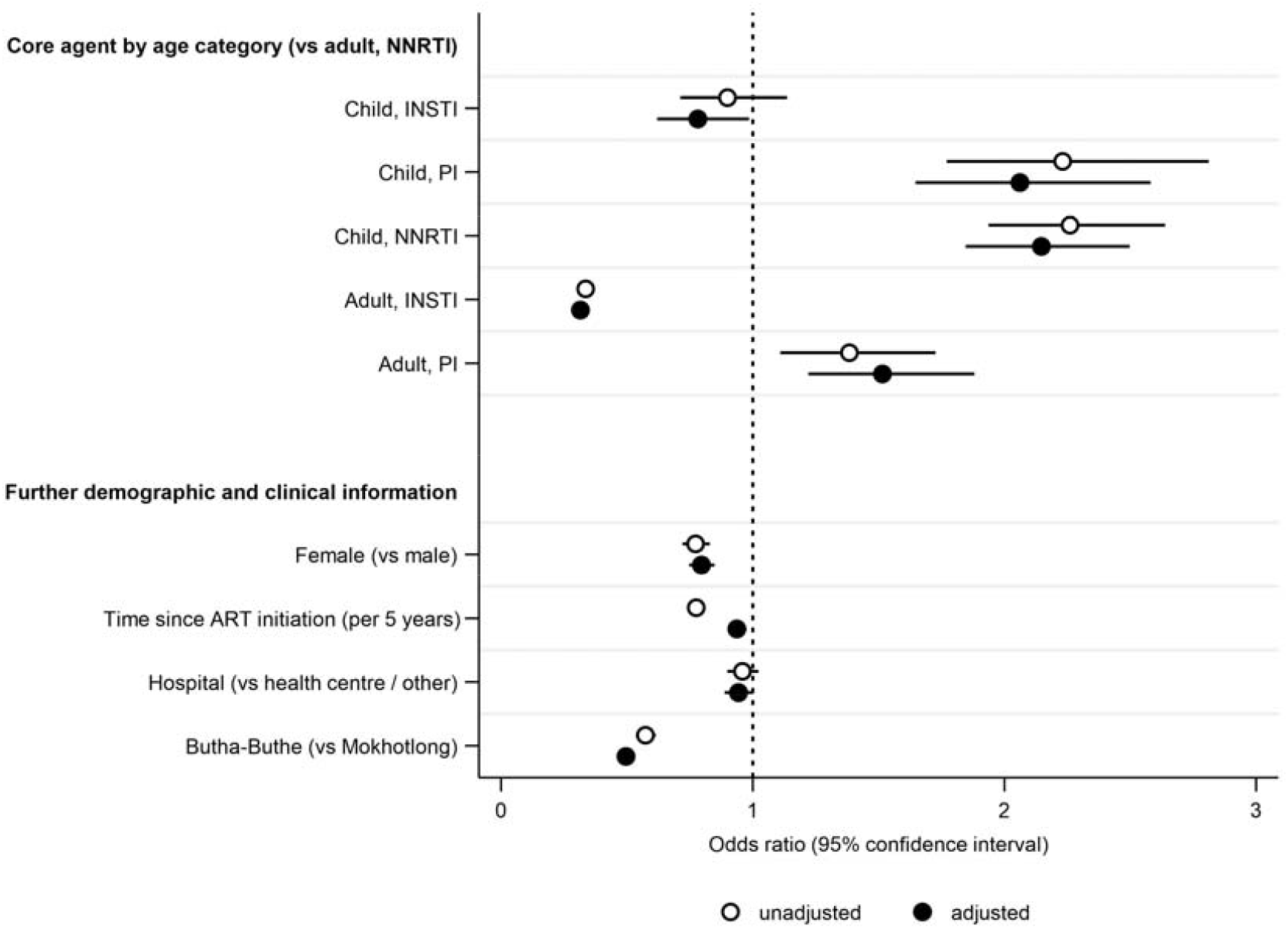
Factors associated with a viral load result ≥50 copies/mL in a logistic regression with mixed effect on the participant level (N=105,183). Odds ratios and 95% confidence intervals of a given viral load result being ≥50 copies/mL are indicated for unadjusted and adjusted analysis. The dotted line at 1 indicates equality of odds. ART: antiretroviral therapy; INSTI: integrase strand transfer inhibitor; PI: protease inhibitor; NNRTI: non-nucleoside reverse transcriptase inhibitor.

Increasing the outcome cut-off to ≥1,000 copies/mL, results were similar except for adults taking PI-based ART, who in this analysis had non-significantly lower odds of viraemia ≥1,000 copies/mL compared with adults taking NNRTI-based ART (**Figure S3**).

### Review of the cohort’s broader findings to date

Key findings from the VICONEL cohort thus far have highlighted gaps in management of HIV viraemia, assessed implications of the rollout of dolutegravir, leveraged the cohort biobank to assess serological profiles during the COVID-19 pandemic, and paved the way for an ongoing nested cluster-randomised controlled trial.

First, management of viraemia – i.e., timely follow-up viral load testing with switching to second-line ART if indicated – was assessed among children[5] and adults[4], demonstrating major gaps in the viral load care cascade. Among children and adults with a viral load ≥1,000 copies/mL, only 28% and 25%, respectively, were considered to be managed in a timely manner according to guidelines.

Second, implications of the large-scale programmatic rollout of dolutegravir-based ART were assessed in children, adolescents, and adults. For adults changing from efavirenz-based ART, we observed no negative impacts of changing to dolutegravir on self-reported symptoms, with potential moderate improvements observed for questions relating to depressed mood, anxiety, and nightmares or strange or vivid dreams[10]. We found that previously described associations of low-level viraemia (50-999 copies/mL) with subsequent viraemia ≥1000 copies/mL hold true also in the era of dolutegravir, supporting the 2021 change in World Health Organization guidelines lowering the viral load threshold for clinical action[7,21]. Overall, short-[22] and long-term viral load outcomes after changing from NNRTI-to dolutegravir-based ART were encouraging, with >95% of adults and >90% of children and adolescents with non-missing data at 24 months having viral suppression to <50 copies/mL[6,8]. However, our recent findings raise important concerns regarding emergent resistance to dolutegravir: among the few participants with persistent and/or recurring viraemia including at least one viral load ≥500 copies/mL at least 18 months after starting dolutegravir, dolutegravir resistance was observed for approx. 10%[23].

Third, serological responses to endemic human coronaviruses and SARS-CoV-2 during the first 18 months of the COVID-19 pandemic were measured using stored biobank samples. This allowed for more detailed reporting on early pandemic dynamics than had previously been available for Lesotho. We identified associations of female sex and high BMI with SARS-CoV-2 seropositivity, and observed positive correlations between the strength of response to endemic HCoVs and to SARS-CoV-2[11].

Finally, formative research on client preferences[12] helped inform the design of the *VIral load Triggered ART care Lesotho* (VITAL) trial, an ongoing cluster-randomised controlled trial within VICONEL. VITAL aims to reduce waiting times, unburden health centres, and reallocate client and healthcare resources in a needs- and preference-driven manner. To do so, the VITAL intervention entails a viral load-driven differentiated service delivery model with automated SMS-based viral load result reporting to participants and an app-based eHealth clinical decision support tool for ART nurses[13].

## Strengths and limitations

Analyses within VICONEL have several limitations. The database is slim and routinely captures minimal demographic, treatment, and clinical data only, whereas data on clinical visits and pharmacy dispensing are not available. This implies that events such as hospitalisations or death are not routinely captured. As data capture occurs only at the time of viral load testing, retention in care cannot be ascertained for those without continued viral load testing.

However, VICONEL also has several strengths. Covering two districts, it is highly representative of people on ART receiving viral load testing in this setting. Data quality is strengthened through obtaining viral load results directly as instrument exports, and through multi-level data checks. Aggregate reports and dashboards aim to support clinical management. Operationally, the reliance on routine data collection keeps maintenance costs low while allowing for near-real-time monitoring in a resource-limited setting; furthermore, the existing framework and biobank facilitate integration of nested non-routine research. Finally, VICONEL has been a successful project for sustainable capacity-building: initial laboratory infrastructure and operational costs were first largely funded by research grants. Subsequently, the Ministry of Health has gradually taken over costs for instrument maintenance and replacements, reagents and consumables, and partial human resource costs. As of December 2022, operational costs (excluding data management and analysis) are limited to salaries for two laboratory technologists and two data clerks, in addition. As such, VICONEL constitutes a model for integrated cross-sectional implementation research with stepwise handover of responsibilities from research and development actors to the Ministry of Health.

## Future plans

While VICONEL is currently highly representative but slim, there is substantial potential for further integration and thematic expansion. In line with the objective of supporting clinical management, this includes further digitalisation and automation of results sharing with participants, healthcare providers, and policymakers, integrating learnings from the VITAL approach. Thematic expansions may include i) optimising integration of point-of-care viral load testing, ii) integration of HIV genotypic resistance test results, iii) expansion to diagnostic data on co-morbidities including tuberculosis and (risk factors for) cardiovascular diseases, and iv) follow-up of mother-child pairs in line with targets for elimination of vertical transmission[24]. Finally, as the cohort grows in scope, we foresee the potential to embed further randomised trials, using innovative designs such as the trials within cohorts (or cohort multiple randomised controlled trial) approach[25,26].

### Ethics approval

The National Health Research Ethics Committee in Lesotho has approved the cohort study (ID 134-2016) and waived consent for analyses using routine data. A biobank consent form for the further use of plasma left over after viral load testing is periodically offered at several sites. Several studies nested within VICONEL but involving non-routine procedures have required a separate study protocol and separate informed consent; data from such studies is not analysed here except insofar as it consists of additional viral load testing or overlaps with routinely collected data.

### Patient and public involvement

The Ministry of Health of Lesotho and the District Management Teams of Butha-Buthe district were among the initial members of the research consortium. Patients and/or the public were not involved in the design, conduct, analysis, or reporting of this research.

### Data sharing

Aggregated or de-identified individual patient data can be shared upon reasonable request and signing of a data sharing agreement. Investigators must submit a concept sheet detailing the required data and planned analyses to the corresponding author for internal review and approval. Publications arising from VICONEL data are subject to written approval by the Sponsor/Chief Investigator and/or the Principal Investigator. VICONEL investigators or contributors shall be co-authors on publications using VICONEL data, provided they fulfil authorship criteria as defined by the International Committee of Medical Journal Editors.

## Author contributions

NDL, TRG, JM, ML, and TK were central to designing and establishing the viral load laboratory, database, and cohort. JAB, NT, and AA have contributed significantly to ongoing development of the cohort. Several authors are or have been centrally involved in oversight (notably NDL, JAB, IA, TRG, JM); clinical management (NDL, AA, BL, BLN, IA); laboratory/diagnostics (JAB, TK); data management and statistics (LM, MGC, MoliM, MauK, KH, FC, NT, JAB, TRG), and coordination or operational management or support (MpK, MatK, MoleM, ML, JAB, NT, NDL, LM, IA, JM, JE). JAB analysed the data with support from FC, NT, and NDL and drafted the first version of this manuscript. All authors reviewed and contributed to the manuscript.

## Acknowledgements

The research consortium consists or previously consisted of members of the Ministry of Health of Lesotho, the District Health Management Teams of Butha-Buthe and Mokhotlong districts in Lesotho, Butha-Buthe Government Hospital, Seboche Mission Hospital, the non-profit organisation SolidarMed, the Swiss Tropical and Public Health Institute, the Department of Biomedicine at the University of Basel, and the Division of Clinical Epidemiology at the University Hospital Basel and University of Basel. The cohort database was developed and is maintained by VisibleSolutions. We wish to thank and acknowledge the clinical and laboratory staff at all healthcare facilities in Butha-Buthe and Mokhotlong districts and at SolidarMed. Finally, we gratefully acknowledge the participants.

The authors would further like to acknowledge the contribution of Christiane Fritz, who sadly passed away before publication of this manuscript.

