## Supplement for "Cohort profile: the Viral load Cohort North-East Lesotho (VICONEL) from 2016 to 2022 – cohort description, test volumes, predictors of viraemia, and the road ahead"

### Supplementary Material

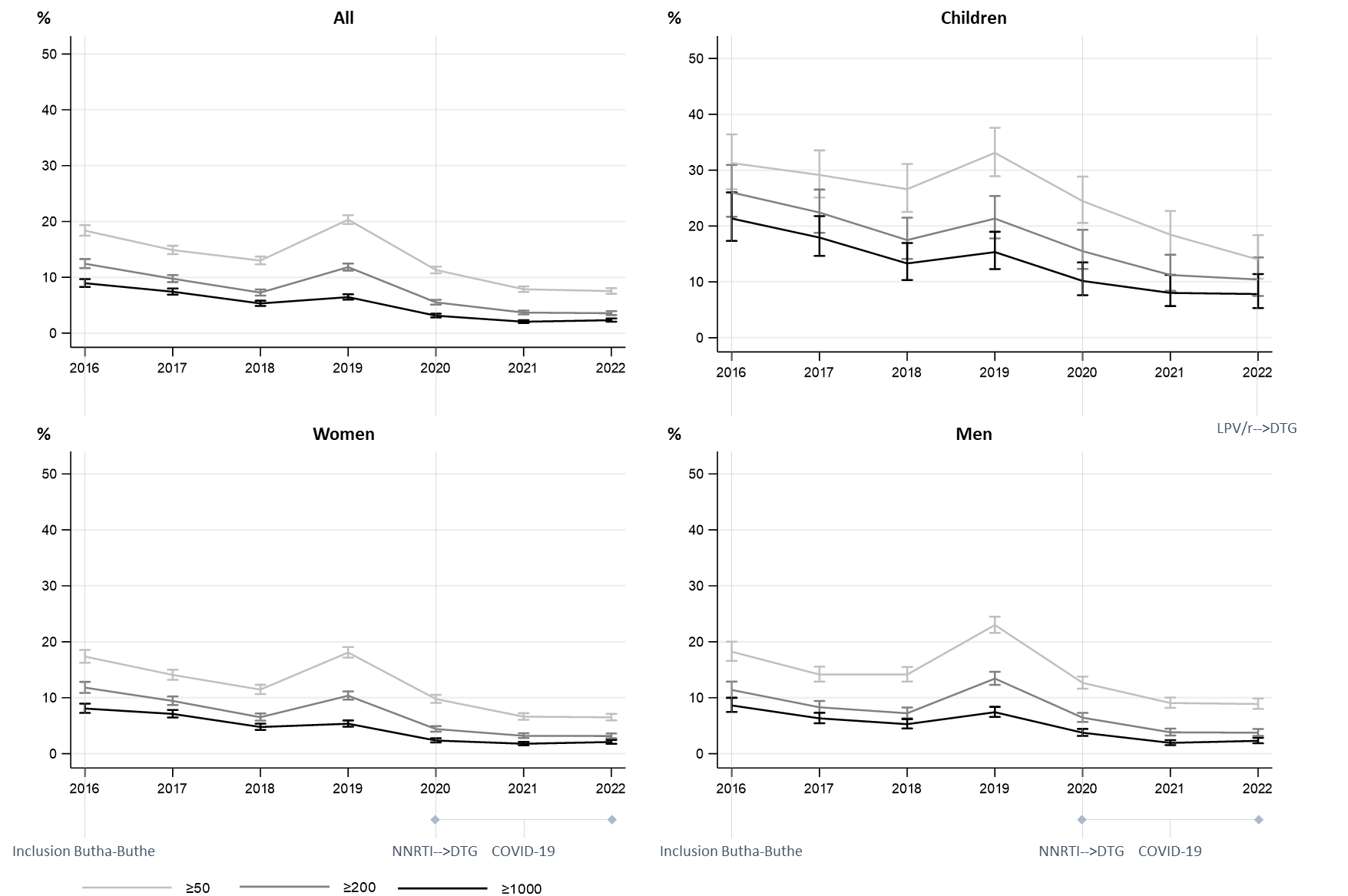

**Figure S1: Rates of viraemia above various thresholds (in copies/mL) over time in Butha-Buthe district.** The first viral load result of any individual in a given year is considered. The denominator corresponds to the number of individuals receiving at least one viral load test in a given year. DTG: dolutegravir; LPV/r: ritonavir-boosted lopinavir; NNRTI: non-nucleoside reverse transcriptase inhibitor.

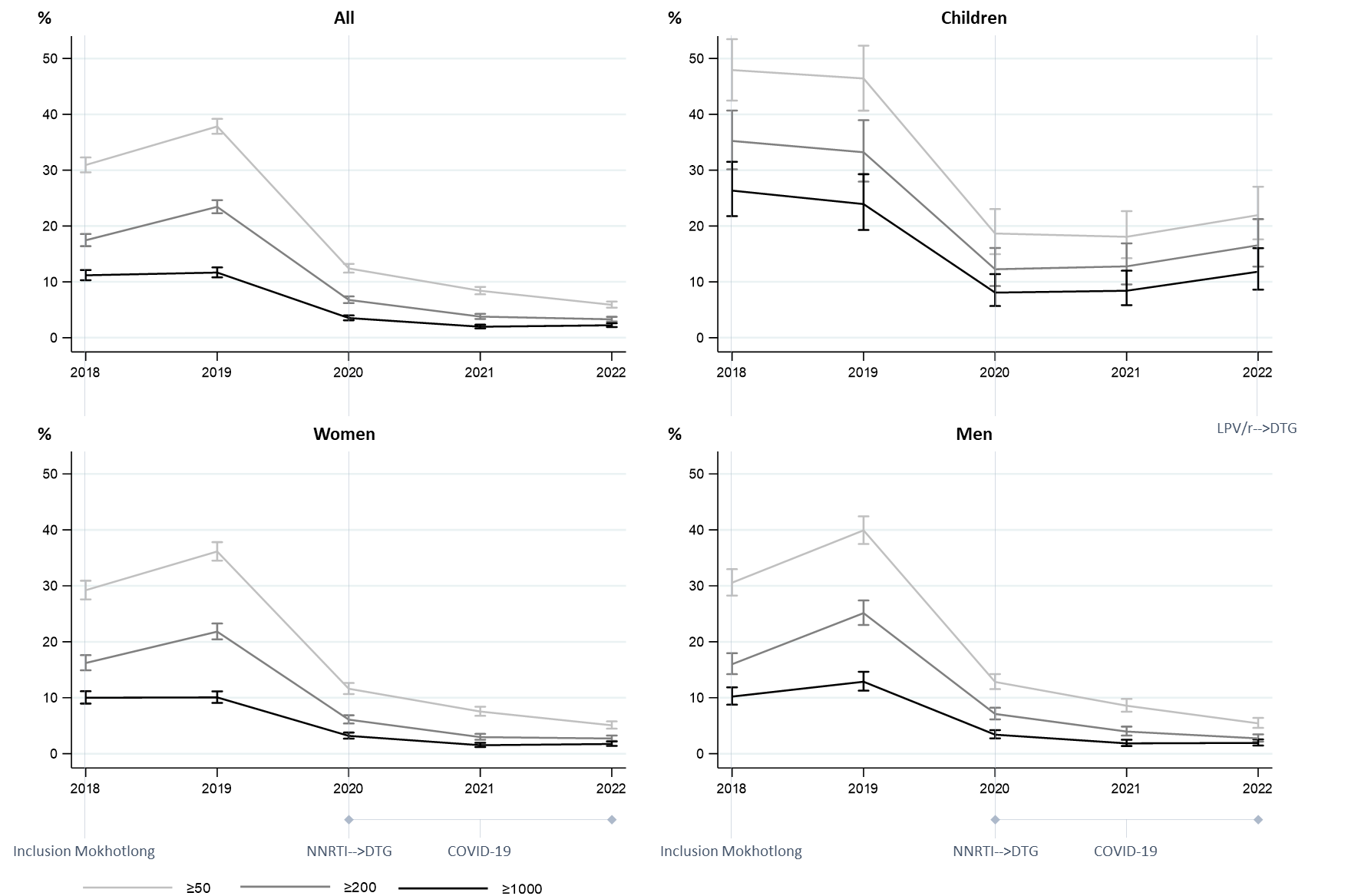

**Figure S2: Rates of viraemia above various thresholds (in copies/mL) over time in Mokhotlong district.** Mokhotlong district became part of the cohort from 2018. The first viral load result of any individual in a given year is considered. The denominator corresponds to the number of individuals receiving at least one viral load test in a given year. DTG: dolutegravir; LPV/r: ritonavir-boosted lopinavir; NNRTI: non-nucleoside reverse transcriptase inhibitor.

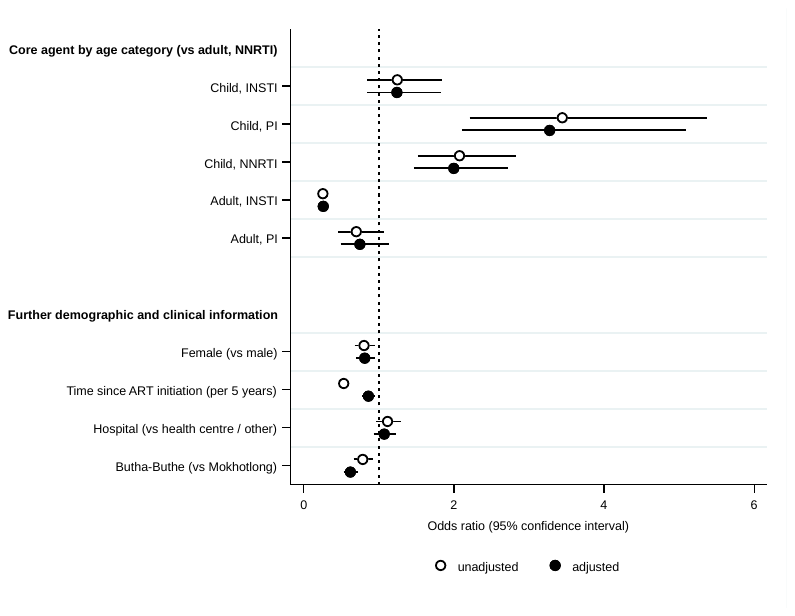

**Figure S3: Factors associated with a viral load result ≥1,000 copies/mL in a logistic regression with mixed effect on the participant level (N=110,844).** Odds ratios and 95% confidence intervals of a given viral load result being ≥1,000 copies/mL are indicated for unadjusted and adjusted analysis. The dotted line at 1 indicates equality of odds. ART: antiretroviral therapy; INSTI: integrase strand transfer inhibitor; PI: protease inhibitor; NNRTI: non-nucleoside reverse transcriptase inhibitor.
